## Supplementary figures and images for "Hip, knee, and ankle joint forces during exoskeletal-assisted walking: comparison of approaches to simulate human-robot interactions"

### Supplemental Figure 1

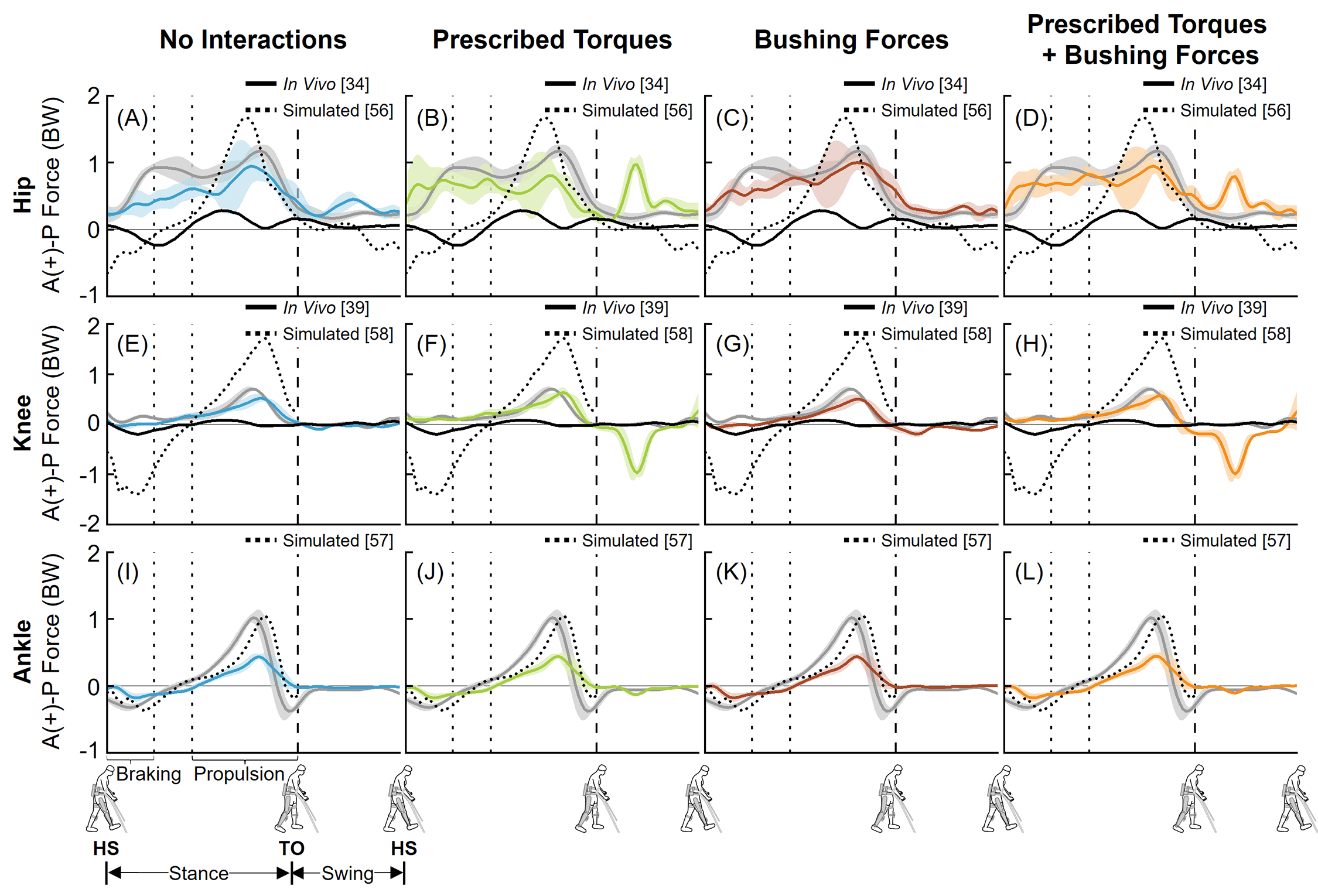

### Supplemental Figure 2

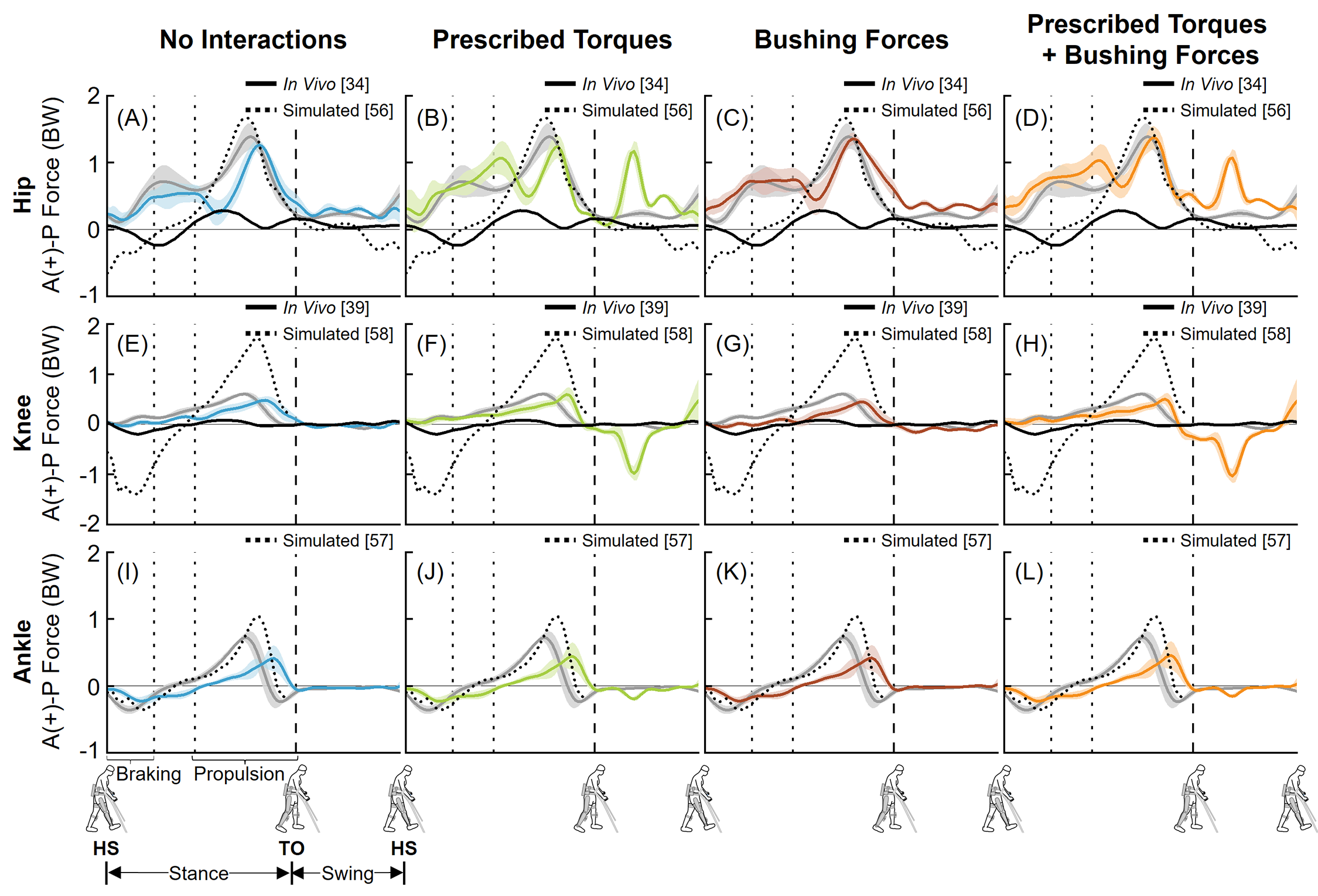

### Supplemental Figure 3

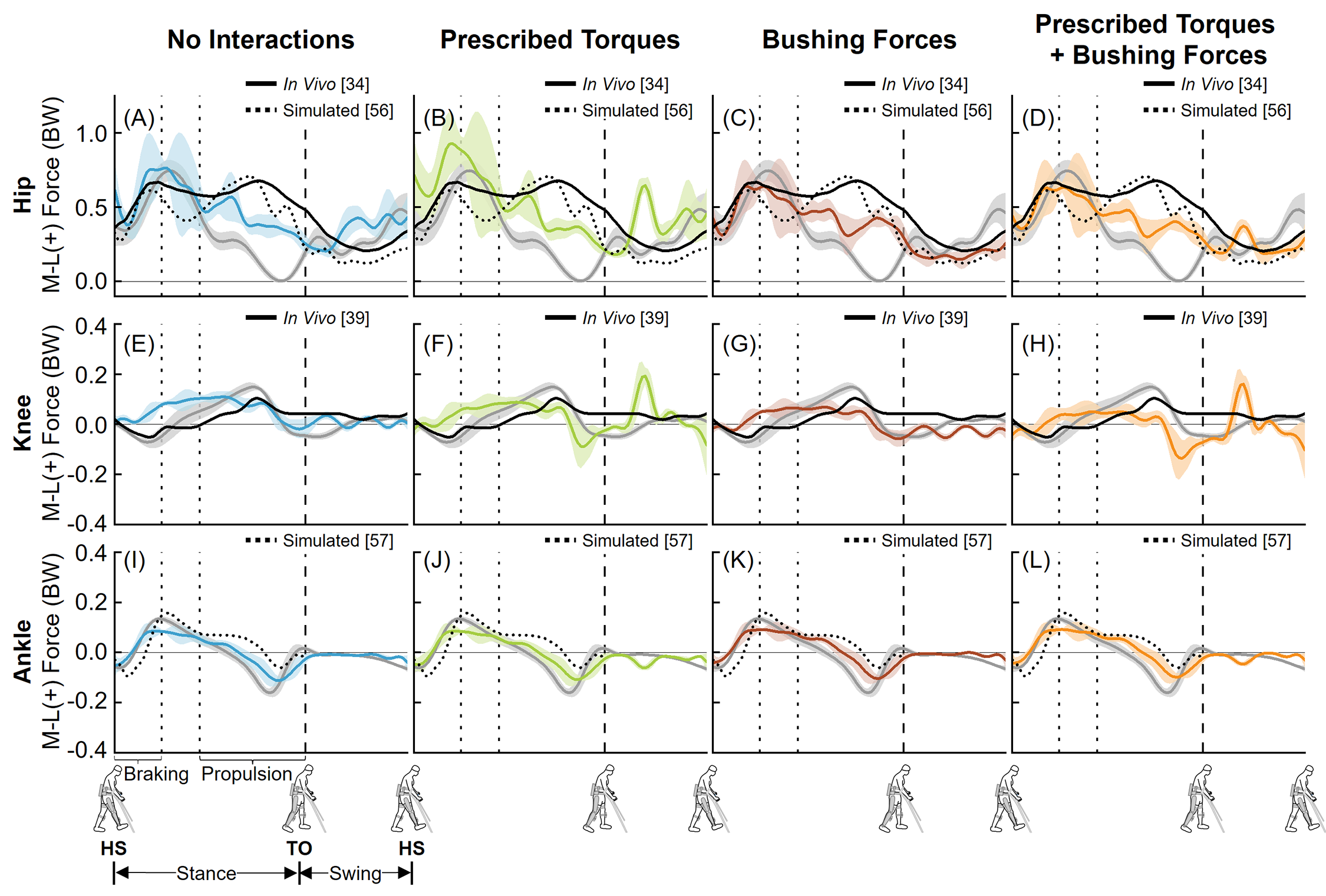

### Supplemental Figure 4

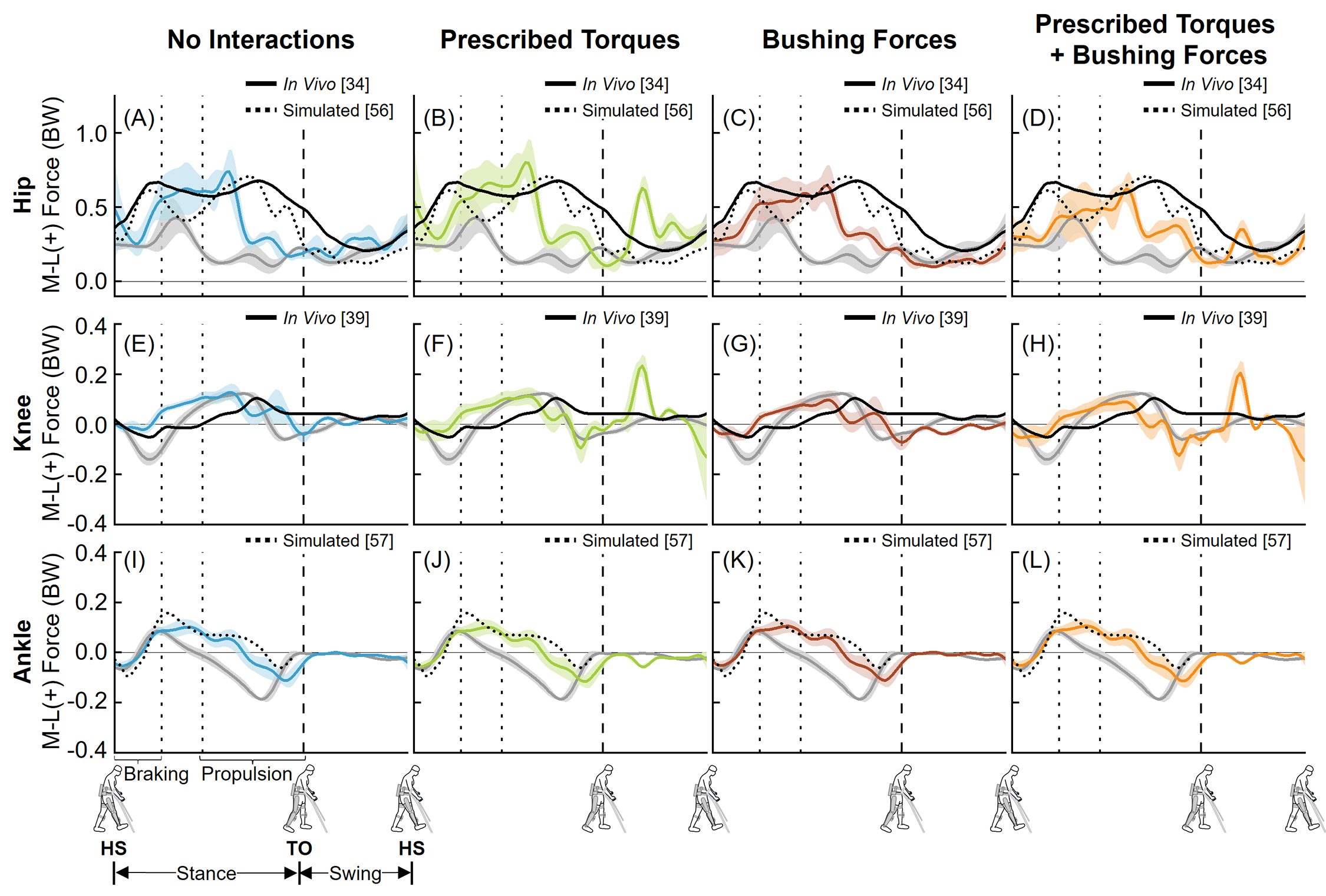
